# SCGAN: Sparse CounterGAN for Counterfactual Explanations in Breast Cancer Prediction

**DOI:** 10.1101/2023.04.16.23288633

**Authors:** Siqiong Zhou, Upala J. Islam, Nicholaus Pfeiffer, Imon Banerjee, Bhavika K. Patel, Ashif S. Iquebal

## Abstract

Imaging phenotypes extracted via radiomics of magnetic resonance imaging have shown great potential in predicting the treatment response in breast cancer patients after administering neoadjuvant systemic therapy (NST). Understanding the causal relationships between the treatment response and Imaging phenotypes, Clinical information, and Molecular (ICM) features are critical in guiding treatment strategies and management plans. Counterfactual explanations provide an interpretable approach to generating causal inference. However, existing approaches are either computationally prohibitive for high dimensional problems, generate unrealistic counterfactuals, or confound the effects of causal features by changing multiple features simultaneously. This paper proposes a new method called Sparse CounteRGAN (SCGAN) for generating counterfactual instances to reveal causal relationships between ICM features and the treatment response after NST. The generative approach learns the distribution of the original instances and, therefore, ensures that the new instances are realistic. We propose dropout training of the discriminator to promote sparsity and introduce a diversity term in the loss function to maximize the distances among generated counterfactuals. We evaluate the proposed method on two publicly available datasets, followed by the breast cancer dataset, and compare their performance with existing methods in the literature. Results show that SCGAN generates sparse and diverse counterfactual instances that also achieve plausibility and feasibility, making it a valuable tool for understanding the causal relationships between ICM features and treatment response.

## 1 Introduction

Breast cancer screening and diagnosis frequently employ Magnetic Resonance Imaging (MRI), a standard of care imaging technique to identify patients at the risk of developing breast cancer and assess the extent of cancer after initial diagnosis [1]. With the emergence of radiomics and machine learning, such imaging techniques are increasingly used in decision-making related to breast cancer treatment [2]. One such application is the use of MRI to predict the response of neoadjuvant systemic therapy (NST) in breast cancer patients. Although NST was initially developed for patients with locally advanced breast cancer, it is now also used for those with early-stage cancer as it may lead to a pathologic complete response (pCR), de-escalating the need for surgery [3]. Accurate prediction of pathologic response to NST could help avoid unnecessary surgical procedures, reduce treatment costs, and minimize side effects [4]. However, determining the suitability of NST for a patient before surgery is complex and depends on various factors, including patient demographics, tumor characteristics, clinical history, and molecular subtypes [5].

Several machine learning models, e.g., random forests [6] and deep learning [7], have emerged to predict the pathologic response of breast cancer patients to NST using pre-operative dynamic contrastenhanced (DCE)-MRI [8, 9]. However, these models do not provide any causal explanations—the cause and effect relationship between features and response—making it difficult to understand the influence of different features such as imaging phenotypes, clinical profile, and molecular makeup (ICM) on the pathologic response. As a result, the wider applicability of these models in clinical decision-making is limited. Initial works focused on generating explanations from black-box machine learning models regarding the most important features in making a prediction, such as LIME (Local Interpretable Modelagnostic Explanations) [10, 11], and Shapley values [12]. Feature importances are generated by recording changes in model’s predictions after adding or removing a feature. However, these models only capture the correlation between features and response, not necessarily their causal relationships [13].

Studying causal inference is important for two main reasons. First, correlation does not imply causation and, therefore, cannot provide a basis for decision-making [14]. Relying solely on correlation without understanding causal relationships can lead to incorrect treatment choices. For instance, several studies have found a positive correlation between alcohol consumption and the incidence of breast cancer [15,16]. However, no conclusive evidence exists to support causal relationship between low to moderate levels of drinking and increased risk of breast cancer [17]. In such cases where no causal relationships exist, correlations may emerge from hidden confounding factors. In the foregoing example, the correlation between alcohol consumption and increased risk of breast cancer may arise from confounding factors such as an indigent lifestyle, family history of breast cancer, or germline BRCA mutation, all of which have been shown to increase the likelihood of developing breast cancer [18]. Second, understanding the causal relationships between ICM features and pCR after NST may lead to opportunities for targeted therapies and help oncologists make informed decisions about continuing or limiting systemic therapy after initial consultation [19].

In a recent publication, Pearl et al. argued that counterfactual explanations could provide the highest level of interpretability in machine learning models and serve as a basis for generating causal inferences [20]. Counterfactual explanations attempt to answer the question about an alternate reality. For instance, “What would have happened if factor X had been observed differently?” In other words, they provide an explanation of how the outcome of an event or decision might have changed if some of the input variables or features had been different [21]. Ideally, counterfactuals should have as few features as possible differently expressed to isolate the effect of causal factors [22]. In the context of predicting treatment response, counterfactual explanations could reveal the set of causal ICM features that would be expressed differently if a patient achieved pCR instead of a pathological non-complete response (pNR).

Various techniques for generating counterfactual explanations have been developed, including model-agnostic, model-specific, and adversarial methods [23–25]. However, critical challenges exist in ensuring feasibility (or realism), promoting diversity and sparsity, as well as scaling them to high-dimensional problems. For example, Diverse Counterfactual Explanations (DiCE), a model agnostic approach, requires minimizing a loss function to generate counterfactuals for each original instance [23]. Scalability becomes a major challenge when generating multiple counterfactual instances using DiCE, particularly for high-dimensional problems. Furthermore, strict optimization approaches fail to ensure realism, which measures how well the counterfactual fits the data distribution [25].

Generative models such as CounteRGAN (CGAN) overcome the challenges of optimization-based methods by generating samples from the original data distribution, leading to realistic and actionable counterfactuals [25]. By producing samples from the original data distribution, CGAN resolves the scalability issues of traditional optimization methods. However, generative methods, particularly, GAN, suffer from mode collapse where the generator only produces new samples from a narrow space in the original data distribution [26]. As a result, the generated counterfactual instances tend to lack diversity. Generating diverse counterfactuals is critical in identifying all causal features while avoiding redundancy of explanations [13]. CGAN attempts to address some of the challenges linked to mode collapse by using a residual GAN [25]. However, it leads to counterfactuals with slight alterations to multiple features, which may not be effective for controlling sparsity. As noted earlier, controlling the sparsity of the counterfactuals is crucial in isolating the effect of different causal features [22].

The current paper builds on our initial work on generating causal inference via counterfactual explanations to predict breast cancer treatment response from ICM features [27]. Our initial work employed DiCE to generate counterfactuals. However, due to the computational cost of DiCE, we preselected the top 10 features using Shapley values [27]. Limiting the feature space significantly restricted the ability of our method to investigate the original domain that comprised 536 features (see Section 4.3). This paper overcomes the aforementioned limitations and extends our initial work by proposing a generative approach called Sparse CounteRGAN (SCGAN) to generate sparse, feasible, plausible, and diverse counterfactuals while keeping computational costs low. In particular, our work improves upon CGAN by constraining the generator to produce sparse and diverse counterfactuals. We accomplish sparsity via dropout training of the discriminator and promote diversity by maximizing the distance among the generated counterfactuals while minimizing their distance from the original instances to maintain proximity. Furthermore, we integrate a masking approach to protect immutable features from changing, thereby ensuring plausibility. Through numerical experiments and real case studies, we demonstrate the efficacy of our approach over existing methods, such as DiCE and CGAN, in generating sparse counterfactual instances with minimal feature changes while maintaining feasibility and enhancing diversity.

The rest of the paper is organized as follows. Section 2 introduces the concept of counterfactuals and the existing counterfactual generation models. Section 3 explains the details of our proposed counterfactual generation approach SCGAN. Section 4 presents the experimental results of SCGAN on two benchmarks and applies it to a realistic breast cancer dataset, comparing its performance with two existing counterfactual generation methods. We use SCGAN to extract causal relationships between breast cancer treatment response and ICM features. Finally, Section 5 summarizes our contributions and provides directions for future research.

## 2 Background and Related Work

Counterfactual refers to a statement or situation describing an alternative outcome or event that could have occurred but did not actually happen [21, 28]. The purpose of generating counterfactual instances in machine learning is to explore what-if scenarios, thereby revealing the causal relationship between input features and response. In particular, what-if scenarios provide insight into how changing certain features might affect the response by generating counterfactual instances and, thereby, finding the hidden causal links.

The origin of counterfactual explanations can be traced back to Hume’s philosophy, which proposed that causation involves an event followed by another, where the absence of the first would prevent the second [29]. Later, this idea gained more prominence through the works of Lewis [30]. In recent years, different approaches for generating counterfactual explanations have been proposed, such as random feature permutation [23, 31], constraint satisfaction [32], mutation and crossover methods inspired by genetic algorithms [33], probabilistic models [34], as well as methods that incorporate LIME and SHAP to perturb features that are the most critical in explaining black-box models [35, 36]. In the following, we first provide a list of properties we seek in counterfactuals and subsequently provide an account of the prominent techniques for generating them.

Chou et. al proposed a set of properties desired in counterfactual instances to ensure meaningful explanations. These are proximity, plausibility, sparsity, diversity, and feasibility in generating counterfactuals [13]. *Proximity* measures the distance between the original instance and the counterfactual [37]. This metric helps assess how much the counterfactual deviates from the original instance. Counterfactuals with lower proximity to the actual instances are sought after. *Plausibility* ensures that the generated counterfactuals are valid and logically reasonable [37]. For instance, a desirable counterfactual should avoid changing immutable features such as gender or race, as these changes are not plausible in real-world scenarios and may have an inherent bias. For example, explanations like “if a patient were caucasian, they would respond positively to the treatment.” Such counterfactuals introduce bias towards the general population and are infeasible in the real world. *Sparsity* pertains to efficiently determining the minimum set of features that must be modified to produce a counterfactual [38]. By changing as few features as possible, the resulting counterfactuals are more interpretable and effective in generating causal explanations by isolating confounding factors. *Diversity* focuses on generating counterfactuals that involve changing different sets of features, thereby creating a wide range of possible explanations. By generating multiple possible explanations, we can identify different causal factors [23,32]. Diversity also promotes interpretability and supports comprehensive explanations for users by providing a thorough understanding of how different factors affect the outcome. Lastly, *feasibility* encourages counterfactuals to be realizable in the real world while staying close to the original instances [39]. For example, counterfactuals like “if the annual wage of a patient (from a socioeconomically underserved population) is doubled, they would have a higher survival rate post-surgery.” While it is possible to change the wage, it might not be feasible without taking other actions, such as earning a degree or learning new skills. Note that the difference between plausibility and feasibility is that the former checks for the correctness of the counterfactuals while the latter is concerned with their practicality.

While an ideal counterfactual should possess all the five properties described in the foregoing, we found that most existing algorithms only achieve some of them. For instance, WatcherCF, one of the first algorithms for generating model-agnostic counterfactuals, only considers proximity and sparsity terms in its loss function [31]. DiCE, an improvement of WatcherCF, includes an additional diversity term in the loss function and incorporates plausibility by adding a constraint for immutable features [23]. However, DiCE does not consider any constraint to check for the feasibility of the generated counterfactuals. In addition to these algorithms, Multi-Objective Counterfactual Explanations (MOCE) proposed by Dandl et al. formulate the counterfactual search as a multi-objective optimization problem using genetic algorithms [33]. However, MOCE does not guarantee plausibility or feasibility. There are other algorithms, such as Model-Agnostic Counterfactual Explanations (MACE) [40] and Recourse [41], that incorporate properties by adding constraints or using probabilistic models, but generating sparse and realistic counterfactual instances remains a challenge. A detailed review of counterfactual generation can be found in [13].

Applying counterfactual explanations for causal inference in the medical field is particularly crucial because of the high stakes involved in medical decision-making. These decisions have serious consequences for patients, so clinicians must thoroughly understand the potential outcomes associated with different treatment options. Counterfactual explanations provide causal relationships between various medical factors, helping clinicians to make informed decisions. In our recent work, Zhou et al. proposed a DiCE-based method for identifying causal relationships between imaging phenotypes, clinical profile, molecular features, and treatment response in breast cancer patients [27]. The authors compared their approach to traditional explanation methods, such as LIME and Shapley, and highlighted the advantages of the counterfactual approach. However, due to the computational complexity of DiCE, the authors used Shapley values to filter out the top 10 most important features for further analysis. It is worth noting that, as Fernandez et al. have shown, a high importance weight from Shapley is insufficient for a feature to be part of a counterfactual explanation [42]. Therefore, the use of Shapley for dimensionality reduction should be considered as an alternative heuristic approach to generate counterfactuals in highdimensional problems.

Due to the challenges of generating high-quality counterfactuals that satisfy the aforementioned properties, recent studies have turned to explore the potential of Generative Adversarial Networks, or GANs, as a viable approach [25]. GANs have been widely used in the field of image generation and have shown impressive results in generating realistic images [43]. GANs generate samples from a complex data distribution by training a generator network to produce samples that are similar to the real data. Trained GANs are suitable for generating counterfactuals since they provide an efficient approach for sampling from any data distribution, in this case, the alternate class. By virtue of generating samples from a specified data distribution, GANs inherently satisfy the feasibility property.

Through incorporating the sparsity and proximity properties for producing high-quality counterfactuals, Nemirovsky et al. adopted GANs to propose a new counterfactual generation approach, called CGAN [25]. This framework generates realistic counterfactuals in achieving the intended class while remaining computationally efficient. In addition to the generator and discriminator networks, CGAN also includes a classifier network. The role of the classifier is to provide additional supervision during training by predicting the responses for counterfactuals. In CGAN, a regularization term is used to encourage sparsity and proximity, which is a combination of the L1 norm and the L2 norm. To ensure plausibility, CGAN cancels the perturbations applied to immutable features after generating the counterfactual instances. However, this strategy may not yield true counterfactuals, especially when changes to immutable features are necessary to realize the alternate response, i.e., one or more immutable features are causally related to the response.

Based on the literature review, it is clear that counterfactual generation has gained significant attention for establishing causal relationships. However, existing methods have limitations in generating counterfactuals that simultaneously meet all properties: proximity, plausibility, sparsity, diversity, and feasibility. These limitations hinder the use of corresponding counterfactual explanations in making accurate causal inferences. This paper builds vertically upon the strengths of CGAN in ensuring feasibility and proximity of counterfactuals by overcoming the limitations of sparsity, diversity, and plausibility, therefore achieving all the desired properties.

## 3 Methodology

### 3.1 CounterGAN

The idea behind GANs is to maintain two neural networks, a generator and a discriminator, that are trained together in an adversarial manner [43]. The generator is a fully connected neural network that produces samples from a target distribution. The discriminator is also a fully connected neural network, but its purpose is to distinguish between the samples produced by the generator (referred hereafter as fake) and the real data, akin to a binary classifier. During training, the generator tries to generate data that mimic the real ones, thereby fooling the discriminator. As the generator gets better at producing realistic data during the training process, the discriminator also improves at distinguishing between the real and fake data. The two networks continue to improve in an adversarial manner until they no longer improve. After the training is complete, the generator is used to sample new instances that resemble the input data distribution [43].

Traditional GAN suffers from mode collapse, causing the generator to produce identical or similar outputs irrespective of the input instances, thereby failing to capture the full diversity and distribution of the training samples. CGAN overcomes some of the issues of mode collapse by generating residuals that, when added to the original instance, produce the desired counterfactual instances.

CGAN also employs a classifier (usually a neural network) to predict the class of the candidate instance, thereby ensuring that it belongs to the alternate class. The classifier network is trained separately before training the generator and discriminator. The overall objective function of CGAN is expressed as:

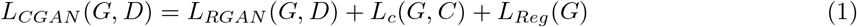

where *G* is the generator network, *D* is the discriminator network, *C* is the classifier network, *L*_*RGAN*_ (*G, D*) is the residual GAN loss, *L*_*c*_(*G, C*) is the classifier loss, and *L*_*Reg*_(*G*) is the regularization term. The Residual GAN loss *L*_*RGAN*_ (*G, D*) in CGAN is formulated as:

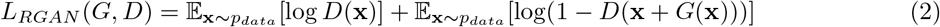

where **x** is an instance from the original dataset, *G*(**x**) is a candidate residual, and **x** + *G*(**x**) is the resulting counterfactual instance. The generator and discriminator are trained simultaneously in an adversarial minimax fashion. The generator network aims to minimize the loss function in Equation (1), while the discriminator network maximizes the same [25].

Without any other constraints, the generator may produce null residuals (i.e., *G*(**x**) = 0) since the resulting counterfactual still remains real and passes the discriminator’s check. The problem of null residuals is avoided by the classifier loss *L*_*c*_(*G, C*) and regularization loss *L*_*Reg*_(*G*) in Equation (1). The classifier loss *L*_*c*_(*G, C*) is formulated as:

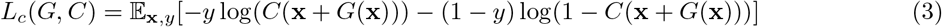

where **x** represents an original instance, *y* is the corresponding label, and *C*(**x** + *G*(**x**)) denotes the output of the classifier network for the candidate counterfactual. The classifier loss measures the binary cross-entropy between the output of the classifier network for the generated counterfactual instance and the opposite label (i.e., 1 − *y*). By minimizing this loss, CGAN ensures that the candidate counterfactual is classified accurately by the classifier network.

The regularization term *L*_*Reg*_(*G*) promotes sparsity and proximity of the counterfactual instances, which is formulated as a combination of L1 and L2 norm, shown as:

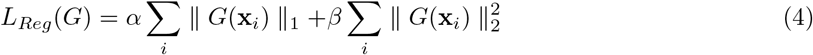

where **x**_*i*_ is an instance drawn from the entire data distribution, *α* ≥ 0 and *β* ≥ 0 are weights of L1 and L2 norm, respectively. Here, the L1 norm encourages the generator to produce residuals where many of the entries are zero, thereby encouraging sparsity. The L2 norm, on the other hand, penalizes the residuals for deviating from zero, encouraging the feature perturbations to be small, but non-zero, thereby promoting proximity.

### 3.2 Sparse CounteRGAN

While CGAN produces counterfactuals that satisfy feasibility and proximity conditions, it fails to ensure sparsity, diversity, and plausibility. First, we investigate sparsity. Even though the L1 norm in Equation (4) promotes sparsity, we note that the generated counterfactuals have small, yet non-zero perturbations to a large number of features. An example can be seen in Figure 2(e) where all the counterfactuals generated by CGAN have perturbation in both the features. The lack of sparsity could be attributed to the training of residual GAN. While generating residual mitigates some of the issues of mode collapse in a traditional GAN, there is no way to enforce the generator to produce residuals from the entire spectrum of the distribution. The L2 norm in Equation (4) simultaneously penalizes the residuals for deviating from zero, encouraging them to be small, yet non-zero. These two factors reduce the sparsity of the counterfactuals.

To overcome the sparsity issue, we remove the L2 norm and adopt dropout strategy in training the discriminator to effectively mitigate the mode collapse issue of residual GAN [26]. The dropout approach involves removing the output of some randomly selected neurons in the discriminator network with a probability *d*, also known as the dropout rate. This causes each neuron to depend on a random subset of the input neurons, thereby capturing the population behavior instead of relying on a fixed set of neurons. Hence, the resulting discriminator is flexible and avoids mode collapse by accepting samples from the entire data distribution. This work considers a dropout rate of 0.9 based on findings in [26].

Second, we look at the diversity of counterfactuals. The existing CGAN architecture does not consider the already existing counterfactuals when creating new ones. To encourage diversity, we introduce a diversity term that penalizes the candidate counterfactuals based on their distance from the already existing ones:

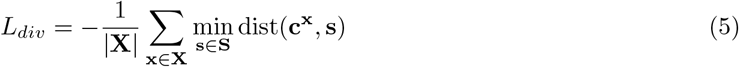

where **X** is the training batch containing the original instances for which we seek to generate counterfactuals, **S** is the set of already existing counterfactual instances, dist(**c**^**x**^, **s**) measures the distance between candidate counterfactual **c**^**x**^ for the original instance **x** and already existing counterfactual **s**. Specifically, the distance dist(**c**^**x**^, **s**) is defined as:

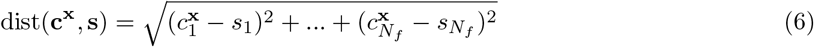

where 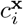 and *s*_*i*_ represent the value of *i*^*th*^ feature in **c**^**x**^ and **s** respectively, and *N*_*f*_ is the number of features. The term min_**s**∈**S**_ dist(**c**^**x**^, **s**) represents the minimal distance between a candidate counterfactual and the already existing counterfactual set. By minimizing Equation (5), we maximize the minimal distance between the counterfactuals. For a new instance for which we have not generated counterfactuals yet, **S** is an empty set. This way, the diversity term penalizes the generator for producing instances similar to the existing ones, thereby producing diverse counterfactuals.

Combining the L1 norm and diversity term, the new regularization loss is expressed as:

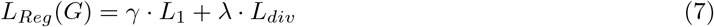

where *γ* ≥ 0 and *λ* ≥ 0 are used to balance the importance of the sparsity and diversity.

Finally, we look at the plausibility of counterfactuals. To ensure plausibility, i.e., avoiding changes to immutable features, CGAN first generates counterfactuals without imposing constraints on immutable features. Any changes observed in immutable features are nullified afterward. However, this may not be ideal, especially when immutable features are causally related to the outcome. In this work, we use a masking approach to restrict the set of features that can be changed in a counterfactual instance. The masking method has been widely applied in image segmentation [44] and natural language processing [45]. In SCGAN, the masking method creates a vector mask consisting of binary values that indicates which features can be changed and which ones cannot. The masking method is applied to the residual part of the generator.

By creating a binary mask, we specify which features are mutable and which are immutable, therefore, ensuring the generation of plausible counterfactual instances. We select the mutable features during training and set their mask values to 1. The remaining features are assigned 0 in the mask. Applying the mask during training ensures that only the mutable features are modified.

In addition to the generator and discriminator, SCGAN also has a classifier, akin to the CGAN model. In binary classification problems, the output of the classifier is a probability value indicating the likelihood of an instance belonging to one of the two classes. However, in the counterfactual generation, it is necessary to make a binary decision based on this probability. This decision is made by applying a threshold to the class probability. By default, the threshold is set to 0.5 in this paper. Figure 1 shows the complete architecture of the proposed SCGAN approach.

**Figure 1:**
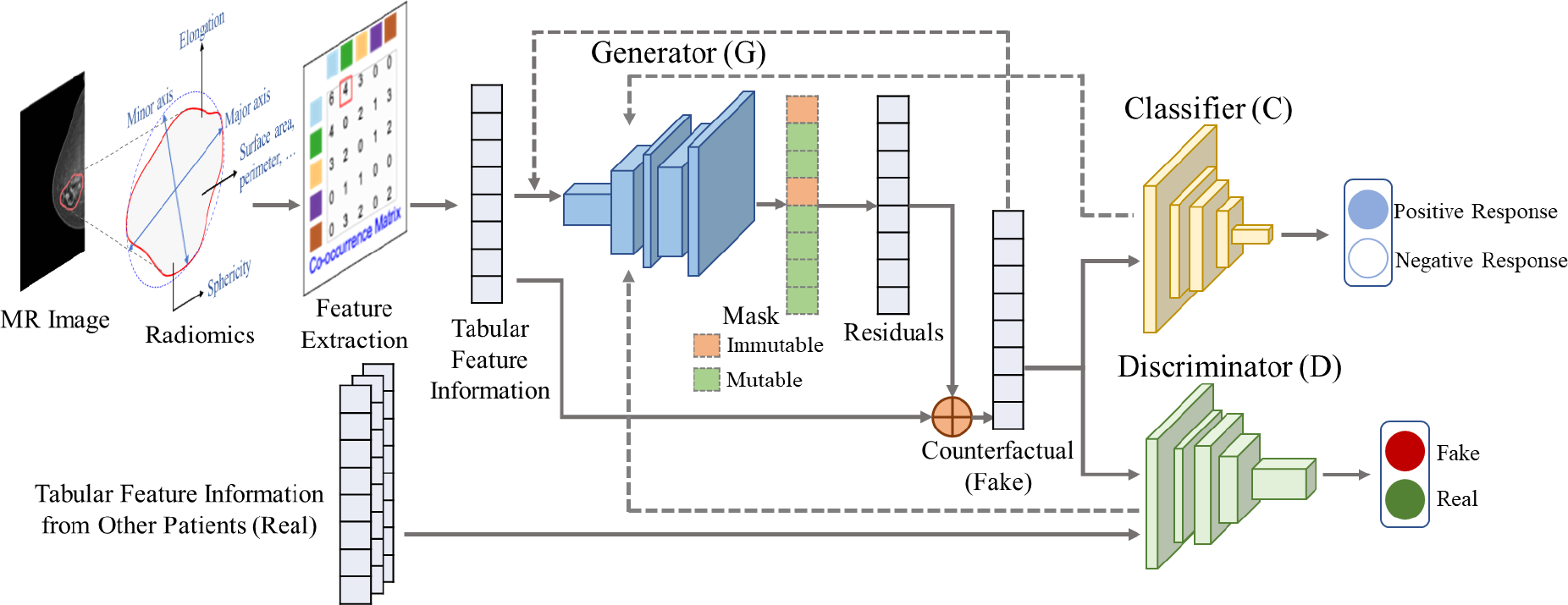
The SCGAN method on an example of breast cancer MR image. The MR image is segmented to identify specific structures of interest such as the breast tissue and tumor. We employ the radiomic features extracted from the MR image to create counterfactual instances [46]. The extracted features serve as the input for training the generator and the discriminator. The generator produces masked residuals that, when added to the input, result in a realistic counterfactual instance. The produced counterfactual instance is then fed back to the generator to encourage diversity. The classifier simultaneously provides feedback on whether the counterfactuals have the desired class.

### 3.3 Evaluation Metrics

We evaluate our method using four metrics. First, we investigate the percentage of generated instances (CF%) that are classified correctly in the alternate class. We accept a generated instance as a counterfactual if the classifier probability exceeds 0.5. Second, we evaluate the average classifier prediction probability (Avg Pred) of the generated instances. A larger value of Avg Pred indicates higher confidence in the classifier’s prediction. Third, we measure the average sparsity (Avg Spar) by counting the number of features changed in the generated instance and averaging over all counterfactuals. The sparsity of a generated counterfactual **c** corresponding to an original instance **x** is defined as:

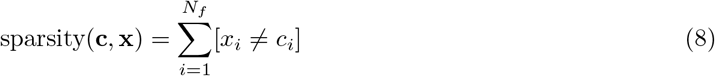

Fourth, we measure average proximity (Avg Prox), which is the average distance between the original instance and the generated counterfactual. Specifically, we use the Euclidean distance d(**c, x**) shown below to measure the distance between an original instance **x** and a generated counterfactual **c**:

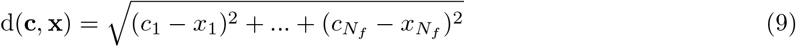

For CF% and Avg Pred, higher values indicate better performance, while for Avg Spar and Avg Prox metrics, lower values are desirable.

## 4 Numerical Experiments

We conduct a comprehensive evaluation of our approach on three distinct datasets: the Pima Indians Diabetes dataset [47] and the Ionosphere dataset [48] as benchmarks, and the Breast Cancer DCEMRI dataset [46] as a case study. We compare SCGAN with two existing counterfactual generation methods, DiCE [23] and CGAN [25]. We utilize a neural network binary classifier to predict the class of generated counterfactual instances [49]. The classifier is pre-trained on the original dataset to achieve a good classification performance. Among various classification methods, neural network delivers the best results. For a comprehensive comparison of different classifiers, please refer to the detailed study presented in [27].

We first evaluate our method on the Pima Indians Diabetes dataset [47], following the experiment in [25]. This dataset comprises of eight features that describe relevant patient characteristics for predicting the presence of diabetes. The classification label is positive if the patient has diabetes (268 instances) and negative otherwise (500 instances). The second dataset we use to evaluate our method is Ionosphere, which has 34 numerical features and binary classification labels for radar returns from the ionosphere [48]. “Good” radar returns (225 instances) are those showing evidence of specific types of structure in the ionosphere. “Bad” returns (126 instances) are those that do not show any evidence. We finally apply our method to the open-source pre-operative DCE-MRI data from Saha et al., which extracted 529 radiomic features for 922 breast cancer patients diagnosed with invasive breast cancer between January 2000 to March 2014 [46]. Please refer to [27] for details on the radiomic features. Out of the 922 patients, 288 were evaluated for neoadjuvant therapy outcomes, categorized as either pCR (64 patients) or pNR (224 patients). To handle imbalance in the breast cancer dataset, random undersampling is applied as discussed in [27].

For the numerical experiments, we employ stratified sampling to split the dataset into training and test sets, guaranteeing a balanced distribution of the target label. This allocation assigned 80% of the data to the training set, leaving 20% for the test set. Table 1 shows the classifier performance across all four cases (two benchmarks and the breast cancer dataset with two different feature sets) for test data. For each dataset, we ensured the model was trained from scratch, allowing it to learn dataset-specific characteristics. Additionally, we employed 5-fold cross-validation based on the specific dataset under consideration to determine the optimal parameter settings. This approach allowed us to select parameters that generalized well to different datasets, ensuring the robustness of our method across various scenarios. All experiments are conducted on a DELL Precision 7865 Tower with AMD Ryzen Threadripper PRO 5945WX 12-core processor and 128 GB RAM.

**Table 1:** Comparison of Classifier Performance.

| Dataset | Accuracy | Precision | Recall | ROC-AUC |
| --- | --- | --- | --- | --- |
| Diabetes | 83.33% | 78.95% | 88.24% | 89.06% |
| Ionosphere | 92.96% | 92.31% | 88.89% | 97.90% |
| Breast Cancer | 57.69% | 69.23% | 56.25% | 60.00% |
| Breast Cancer<br>-Reduced | 76.92% | 80.00% | 66.67% | 77.98% |

### 4.1 Benchmark 1 Pima Indians Diabetes

We first visually illustrate the generation of counterfactual instances with DiCE, CGAN, and SCGAN. Figure 2(a) shows a two-feature projection scatter plots between glucose concentration (*x*-axis) and BMI (*y*-axis) measurements for the Pima Indians. Figure 2(b) shows the decision boundary of the binary classifier. We select four instances (shown in yellow in Figure 2(c)) representing patients without diabetes to generate the corresponding counterfactuals. The counterfactuals generated by DiCE (green dots shown in Figure 2(d)) require a significant change in both glucose and BMI. We also notice that some of the counterfactuals lie near the tail of the original data distribution, making them less likely to be realistic. Using CGAN (as shown in Figure 2(e)), the generated counterfactuals are more realistic and require smaller changes to features, but necessitate altering both features in all four cases. The proposed SCGAN method (as seen in Figure 2(f)) results in counterfactuals that achieve the desired classification and only require altering one feature in two out of the four examples. The resulting counterfactuals are sparse compared to the ones generated by DiCE or CGAN.

**Figure 2:**
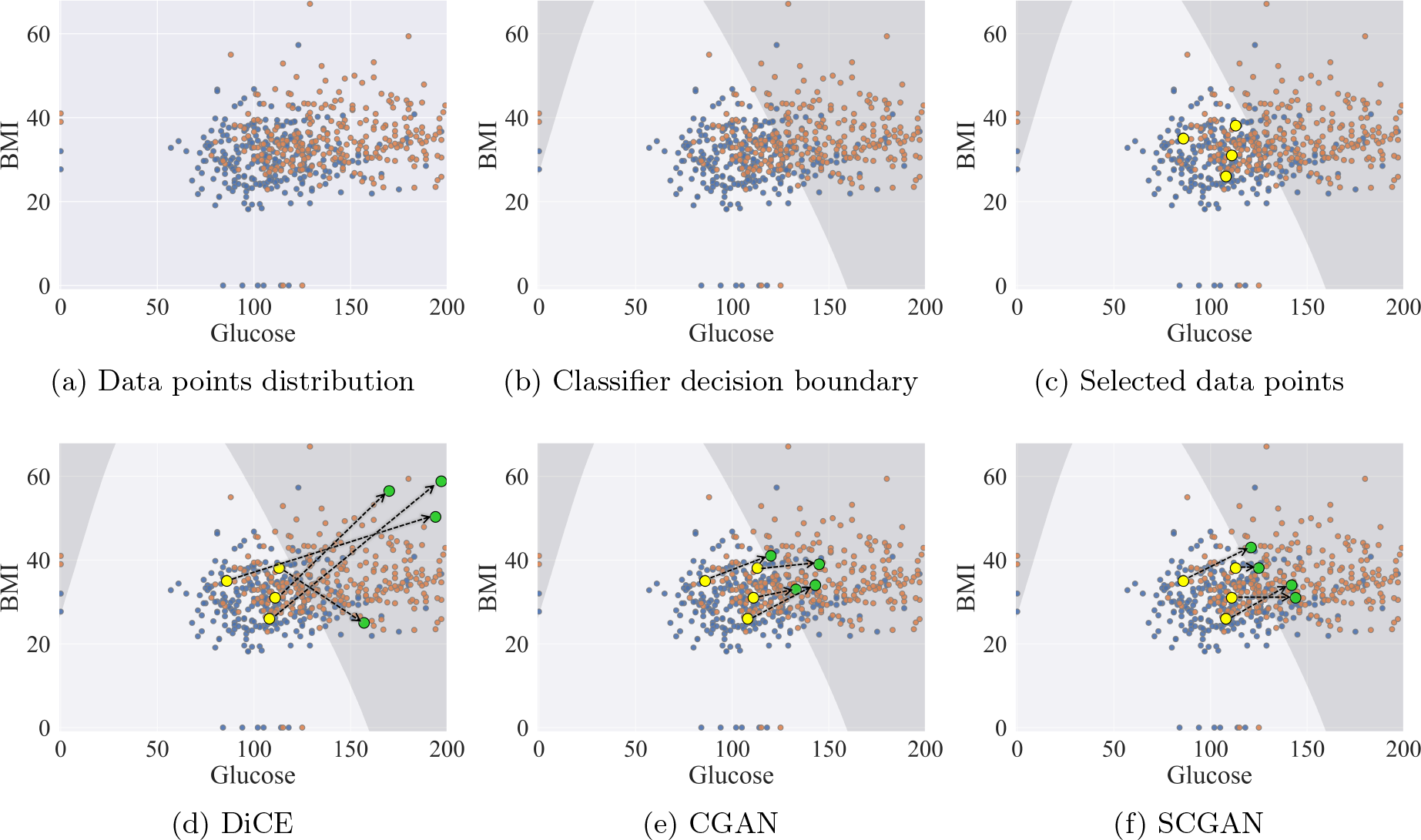
Comparison of three counterfactual search techniques on the Pima Indians Diabetes dataset, showing how they achieve their objectives while generating markedly different counterfactual instances. Due to the two-feature projection, Figure 2 cannot fully capture the counterfactual’s prediction probability level, and the distance to the decision boundary does not necessarily imply the classifier probability in individual predictions.

For the Pima Indians Diabetes dataset, we consider three features, Pregnancies, Age, and Diabetes Pedigree Function, as immutable, while Glucose, Insulin, Body Mass Index, Tricept Skin Fold Thickness, and Blood Pressure are treated as mutable, as suggested in [25]. In this experiment, we compare three methods using randomly selected original instances where the patients with no diabetes are classified as “negative” (class 0). Our objective is to generate instances that belong to the “positive” class (class 1). We generate 50 counterfactuals for each of the five initial instances and summarize the performance in Table 2.

**Table 2:** Model Performance Comparison for Benchmark 1.

| Method | CF% ↑ | | Avg Pred ↑ | | Avg Spar ↓ | | Avg Prox ↓ | | Total Time<br>($5 \times 50$ CFs) |
| --- | --- | --- | --- | --- | --- | --- | --- | --- | --- |
|  | Mean | SD | Mean | SD | Mean | SD | Mean | SD |  |
| DiCE | 96.8 | 2.71 | 0.594 | 0.066 | 2.112 | 0.635 | 0.481 | 0.211 | 15.1 |
| CGAN $\alpha = 1, \beta = 0$ | 66.0 | 28.17 | 0.576 | 0.126 | 1.648 | 0.424 | 0.128 | 0.054 | 5887.9 |
| SCGAN $\gamma = 1, \lambda = 5$ | 92.0 | 14.03 | 0.652 | 0.124 | 1.620 | 0.661 | 0.144 | 0.042 | 3556.9 |
| SCGAN $\gamma = 0.5, \lambda = 5$ | 100.0 | 0.00 | 0.826 | 0.077 | 2.880 | 1.050 | 0.172 | 0.028 | 3640.0 |

All three methods generated counterfactuals with opposite classifier predictions for most instances. SCGAN with *γ* = 0.5 generates counterfactuals for 100% of the instances, surpassing both DiCE (96%) and CGAN (92%). In terms of the average prediction probability, SCGAN generated counterfactuals with higher values than both DiCE and CGAN. Regarding average sparsity, SCGAN with *γ* = 1 achieves the lowest value of 1.62. For the average proximity, SCGAN with *γ* = 1 achieves better performance than DiCE, but slightly worse (around 10%) than CGAN. When comparing the last two rows with different *γ* values, we observe that a smaller *γ* leads to worse sparsity. This is expected since a smaller *γ* imposes a smaller penalty on the number of non-zero perturbations in the features. However, the percentage of generated instances that achieve the opposite class and the average prediction probability of counterfactuals are also higher. While SCGAN is slower as compared to DiCE, run time is not a critical factor in generating counterfactuals. For counterfactual generation, the main priority is producing instances that align with the desired outcomes.

Figure 3 displays the distribution of BMI in the Pima Indians Diabetes dataset (as a histogram) and feature values of generated counterfactuals (vertical lines). We observe that the counterfactual instances generated from SCGAN (blue lines) are more diverse than CGAN (green lines). CGAN and SCGAN generate counterfactual instances with BMI values in a high-likelihood region, making them realistic. On the other hand, the counterfactuals generated via DiCE are diverse but lie near the tail of the original data distribution, suggesting that such instances are less likely to be realistic.

**Figure 3:**
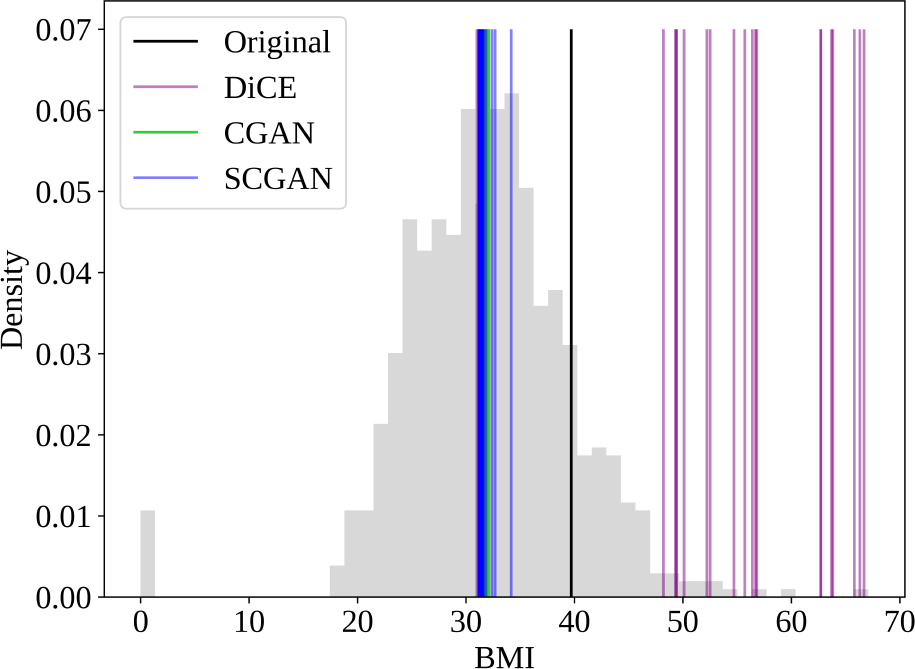
The feature value distribution of BMI in the Pima Indians Diabetes dataset. The histogram shows the distribution of values in the entire dataset. Vertical lines depict the original and counterfactual instances generated by DiCE, CGAN, and SCGAN, color-coded as per the legend.

Figure 4 visualizes the counterfactuals generated by DiCE, CGAN, and SCGAN for a selected initial instance. The counterfactuals are color-coded per their prediction probability, with blue close to 1 (more likely) and red close to 0 (less likely). We see that SCGAN achieves higher prediction probabilities than DiCE and CGAN for most counterfactual instances. SCGAN produces more diverse counterfactuals than CGAN but is not as good as DiCE. However, considering the result shown in Figure 3, SCGAN considers the feasibility requirement, while the counterfactual instances from DiCE may not be realistic as they lie towards the tail of the original data distribution.

**Figure 4:**
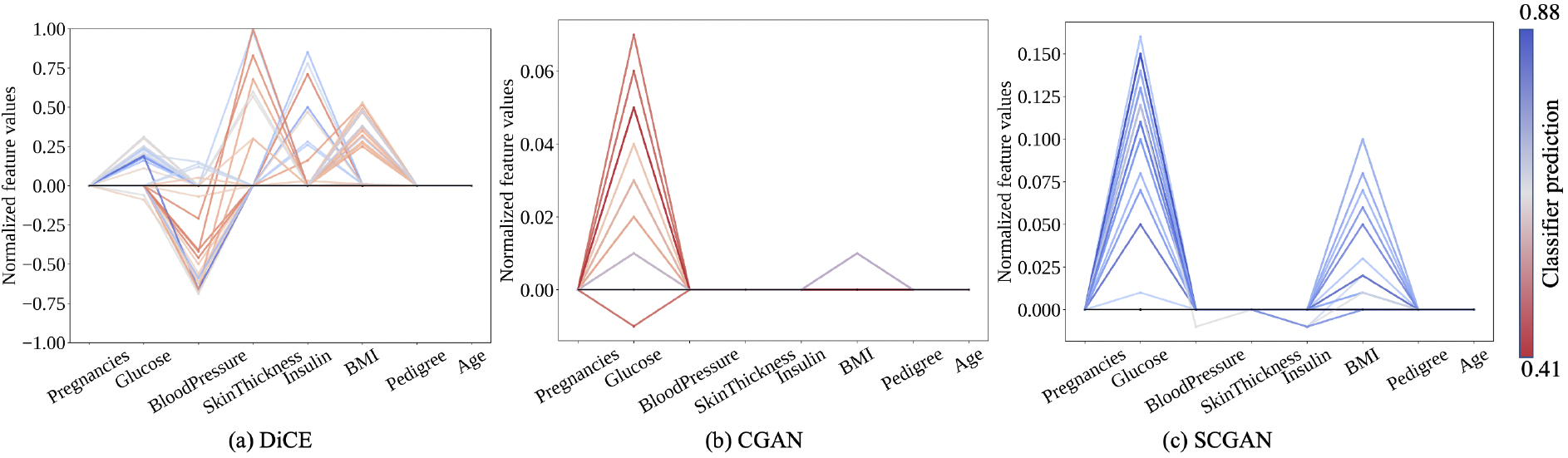
Counterfactual instances generated by three methods. The horizontal axis represents the features and the vertical axis represents the normalized feature values. The black zero line represents the actual instance. Every counterfactual instance has either one or more changes in the feature values, each of which is color-coded based on their classification probability.

Our numerical experiments on the Pima Indians Diabetes dataset show that glucose and BMI are the two most frequently changed features. These results conclude that glucose and BMI are the causal features in diabetes diagnosis. This conclusion aligns with previous findings in [50].

### 4.2 Benchmark 2 Ionosphere

The experimental settings for benchmark 2 are similar to those described for benchmark 1. Table 3 summarizes the three methods’ performance. SCGAN generates instances that achieve the opposite class for most cases (more than 75%), surpassing CGAN (56%) and approaching the performance of DiCE with *γ* = 0.05 (with a difference of approximately 5%). Moreover, SCGAN consistently outperforms the other two methods regarding average prediction probability for both *γ* settings. When evaluating average sparsity, SCGAN with *γ* = 0.5 has a lower value than CGAN and is close to the lowest value achieved by DiCE (with a difference of roughly 8%). For the proximity measurement, SCGAN gets lower values than both CGAN and SCGAN. When comparing SCGAN with different *γ* settings, we observed that a lower *γ* value produces counterfactuals with higher prediction probabilities but compromised sparsity.

**Table 3:** Model Performance Comparison for Benchmark 2.

| Method | CF% ↑ | | Avg Pred ↑ | | Avg Spar ↓ | | Avg Prox ↓ | | Total Time<br>($5 \times 50$ CFs) |
| --- | --- | --- | --- | --- | --- | --- | --- | --- | --- |
|  | Mean | SD | Mean | SD | Mean | SD | Mean | SD |  |
| DiCE | 95.6 | 2.939 | 0.634 | 0.101 | 5.53 | 2.837 | 1.020 | 0.388 | 83 |
| CGAN $\alpha = 0.5, \beta = 0$ | 56.0 | 40.431 | 0.532 | 0.396 | 21.80 | 13.410 | 0.635 | 0.602 | 5154 |
| SCGAN $\gamma = 0.5, \lambda = 5$ | 78.0 | 44.000 | 0.748 | 0.330 | 8.43 | 5.510 | 0.316 | 0.272 | 6845 |
| SCGAN $\gamma = 0.05, \lambda = 5$ | 90.5 | 12.370 | 0.933 | 0.109 | 22.89 | 4.141 | 0.514 | 0.217 | 7157 |

Figure 5 displays the distribution of Attribute 12 in the Ionosphere dataset and the feature values of generated counterfactual instances. We note that SCGAN generates counterfactuals with feature values distributed in a wider range than CGAN, indicating diversity. DiCE is the only method that generates a counterfactual instance with a value above 1, which is beyond the range of the feature distribution in the dataset.

**Figure 5:**
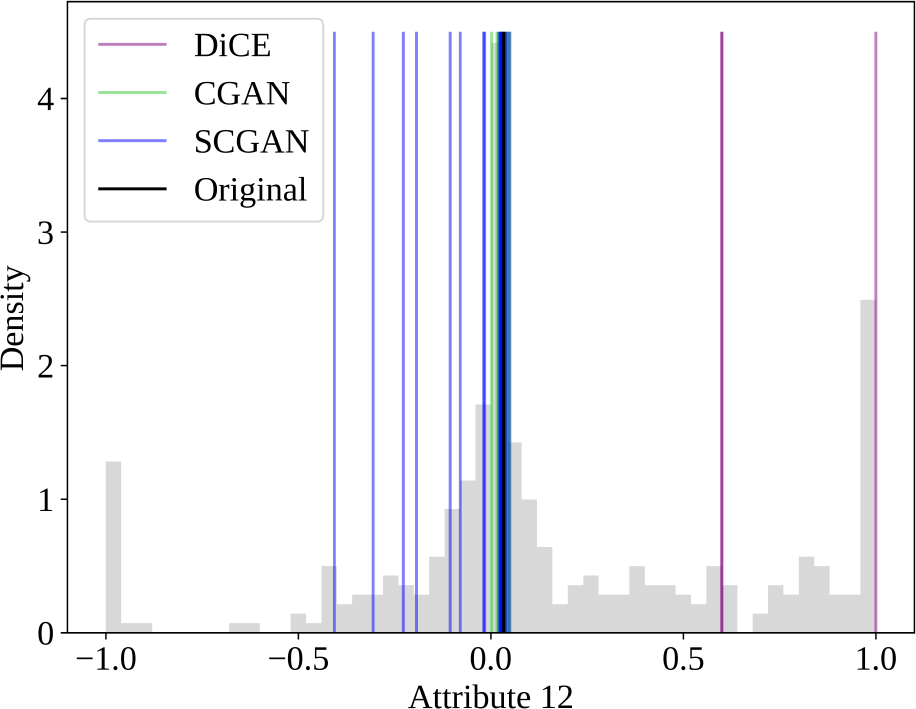
The feature value distribution of Attribute 12 in the Ionosphere dataset. The histogram shows the distribution of values in the entire dataset. Vertical lines depict the original and counterfactual instances generated by DiCE, CGAN, and SCGAN, color-coded as per the legend.

Figure 6 displays the counterfactuals generated by DiCE, CGAN, and SCGAN for a selected initial instance. Carefully observing the figure reveals that SCGAN generates counterfactual instances with smaller changes than DiCE. Compared with SCGAN, CGAN generates instances with smaller changes but also lower prediction probabilities. SCGAN changes the features moderately and guarantees high prediction probabilities and diversity.

**Figure 6:**
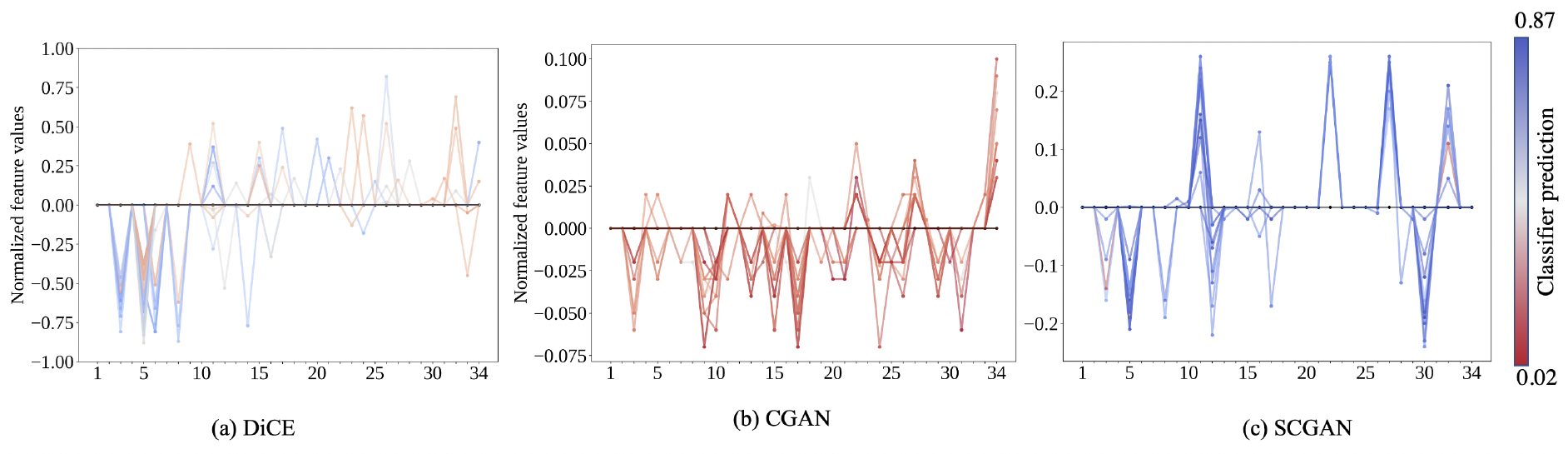
Counterfactual instances generated by three methods. The horizontal axis represents the features and the vertical axis represents the normalized feature values. The black zero line represents the actual instance. Every counterfactual instance is represented as a color-coded line, where the color is based on their classification probability.

The Ionosphere dataset has 34 attributes to describe 17 pulse numbers obtained from the complex electromagnetic signal collected by a system in Goose Bay, Labrador [48]. Based on the results from SCGAN, we observe that the 25th and 27th attributes are the primary contributing factors identifying the “Good” radar signals from the bad ones.

### 4.3 Case Study: Breast Cancer DCE-MRI

We generate 50 counterfactuals using CGAN and SCGAN for each initial instance and summarize the average results in Table 4. SCGAN achieves opposite classifier predictions in more than 90% of instances, surpassing CGAN, which reaches just over 60%. In addition, SCGAN demonstrates higher prediction probabilities in generating counterfactual instances than CGAN. SCGAN also outperforms CGAN regarding sparsity and proximity in both *γ* settings. In particular, a higher *γ* value for SCGAN leads to counterfactual instances with lower prediction probability but fewer feature changes, as shown in the last two rows of Table 4. This trade-off highlights the importance of carefully tuning the regularization coefficient to balance sparsity and prediction probability in counterfactual generation. DiCE cannot generate any counterfactual instances within the time limit of one hour.

**Table 4:**
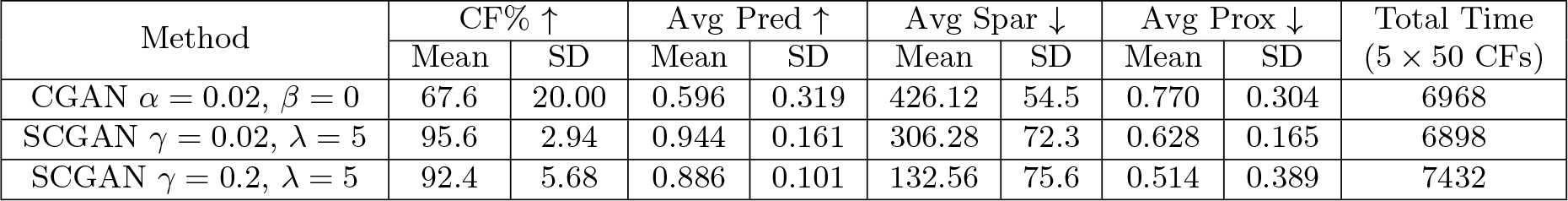
Model Performance Comparison for Breast Cancer DCE-MRI Dataset.

Figure 7 illustrates the distribution of “tumor voxel cluster similarity” (which stands for the similarity of neighboring tumor voxels in the same cluster) in the Breast Cancer dataset and the feature values of generated counterfactual instances. Our proposed method, SCGAN, generates counterfactual instances with feature values that are more diverse and from a higher data density region compared to CGAN. This characteristic of our method demonstrates its ability to generate counterfactual instances that are more realistic.

**Figure 7:**
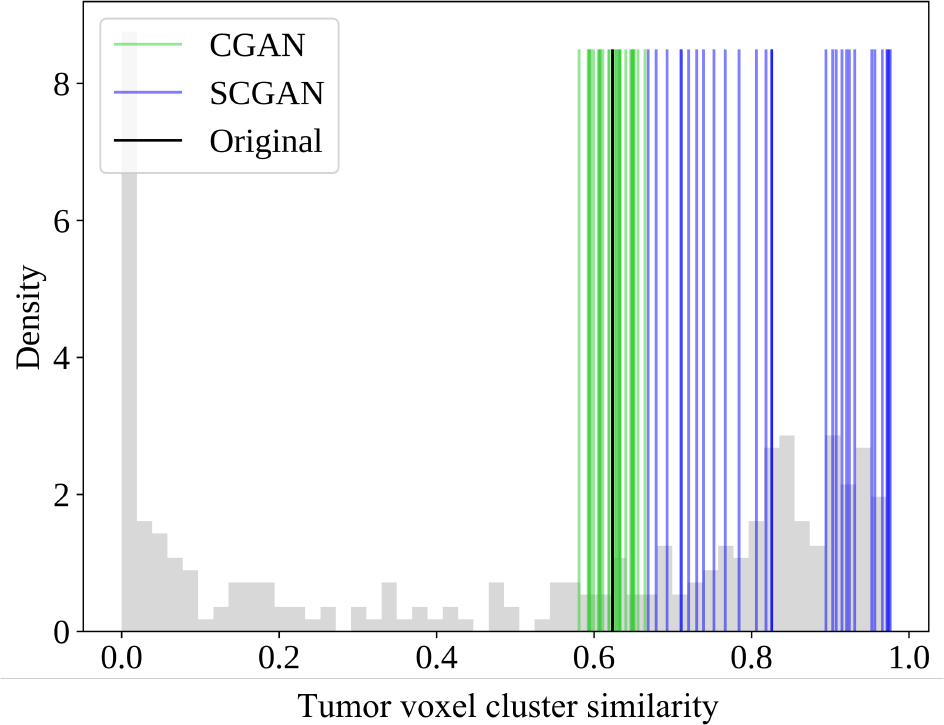
The feature value distribution of the similar clustered tumor voxel proportion in the Breast Cancer dataset. The histogram shows the feature’s value distribution in the dataset. Vertical lines depict the original and counterfactuals generated by CGAN, and SCGAN, color-coded as per the legend.

To avoid confounding effects from correlated features, we filtered out correlated and collinear features. Specifically, we calculate the correlation matrix and look for features with a high correlation coefficient. If a pair of features has a high correlation coefficient (*>* 0.85), then one of them is dropped. Secondly, the variance inflation factor (VIF) is calculated for each feature to assess the degree of collinearity. Features with a high VIF (*>* 10) indicate high collinearity and are dropped. Using these criteria, we filter out 41 features for the subsequent analysis. In this case, even with 41 features, DiCE did not generate any counterfactual instances within the time limit of one hour.

We generate 50 counterfactual instances using CGAN and SCGAN for each initial instance and summarize the average performance in Table 5. Overall, SCGAN produces a higher percentage of instances that achieve the opposite class (CF%) than CGAN. Furthermore, the prediction probabilities of generated counterfactuals from SCGAN are larger. In terms of average sparsity, SCGAN with *γ* = 2 demonstrates the lowest value, approximately 17, among all cases. Even in the case of SCGAN with *γ* = 0.02, the average sparsity is not significantly worse than CGAN (less for around 6%). Based on the average proximity values, we note that SCGAN performs worse as compared to CGAN. This is because categorical features are only allowed to change as integers in SCGAN, but not in CGAN. In the latter, categorical features are treated the same as numerical features and may get decimal values.

**Table 5:** Model Performance Comparison for Reduced Breast Cancer DCE-MRI Dataset.

| Method | CF% ↑ | | Avg Pred ↑ | | Avg Spar ↓ | | Avg Prox ↓ | | Total Time<br>($5 \times 50$ CFs) |
| --- | --- | --- | --- | --- | --- | --- | --- | --- | --- |
|  | Mean | SD | Mean | SD | Mean | SD | Mean | SD |  |
| CGAN $\alpha = 0.02, \beta = 0$ | 66.0 | 23.87 | 0.672 | 0.359 | 27.912 | 1.991 | 0.250 | 0.044 | 2725.3 |
| SCGAN $\gamma = 0.02, \lambda = 5$ | 100.0 | 0.000 | 0.999 | 0.001 | 30.02 | 2.569 | 1.348 | 0.535 | 2763.1 |
| SCGAN $\gamma = 2, \lambda = 5$ | 92.4 | 3.441 | 0.920 | 0.246 | 17.464 | 2.876 | 1.444 | 0.087 | 2919.2 |

Based on the generated counterfactuals from SCGAN, we observe that only three out of the 41 filtered ICM features are involved all the time, namely PR status (which stands for progesterone receptors status), the HER2 status (where HER2 is a protein that helps breast cancer cells to grow rapidly), and the variance of uptake (which captures the inhomogeneity of IV uptake between successive MRI frames), which is consistent with the previous finding in [51]. These are considered causal for predicting the treatment response in breast cancer.

Causal relationships help guide oncologists to optimize decision-making in breast cancer treatment. Healthcare professionals can employ different feasible counterfactual explanations to increase the likelihood of achieving pCR. By emphasizing causal features, they are able to improve treatment outcomes for patients who struggle to achieve pCR. Furthermore, counterfactual explanations identify influential patient-specific features. For instance, if changing PR status consistently leads to pCR for a specific patient, modifying it becomes an option to improve the treatment response. This allows oncologists to apply hormone therapy to block PR-positive tumors before tumor treatment. In our experiment, we removed correlated and collinear features. While this helps eliminate the confounding factors, dealing with correlations in generating causal inference is still challenging. Finding effective ways to handle correlations is important for enhancing the accuracy and applicability of counterfactual generation methods, which in turn will support informed decision-making.

### 4.4 Validating Feasibility of Counterfactuals

As counterfactuals are unobservable in the real world, we utilize a simulated dataset to check the feasibility (or realism) of the results. This controlled environment enables a comparison of the produced counterfactuals against the true instances as specified in the data generation process. To this end, we simulated two groups, each with one feature and 500 instances. The samples in class 0 is generated from *N* (50, 1) and class 1 from *N* (60, 1). We generate counterfactuals for class 0 such that the resulting instances belong to class 1. We compare the distribution of the counterfactuals with the original data in class 1 to assess their feasibility. Figure 8 shows the distribution of the counterfactual instances and the original dataset. We use the two-sample Kolmogorov-Smirnov (KS) test to check if the two distributions are the same [52]. The KS test, conducted at a significance level of 0.05, yielded a p-value of 0.182. This p-value suggests that there is no statistically significant difference between the two distributions. Additionally, visual comparisons of the distributions also support this finding. This simulation demonstrates SCGAN’s ability to produce feasible counterfactuals critical in generating meaningful explanations.

**Figure 8:**
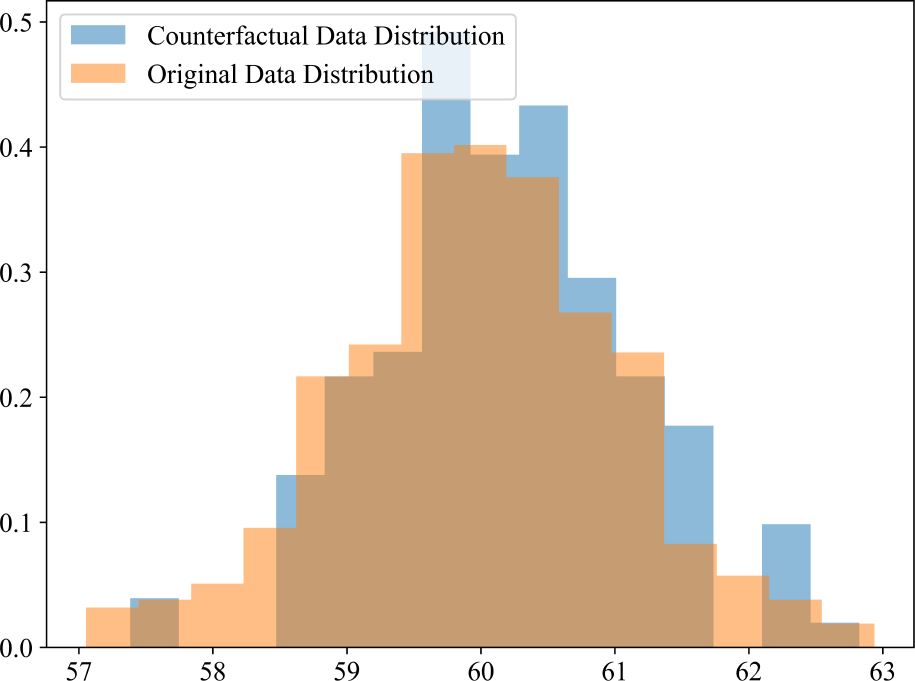
Comparison of distributions for simulated data and generated counterfactuals.

## 5 Conclusions

In this paper, we extend the CGAN approach to generating counterfactual instances that are sparse and diverse to achieve causal inference and overcome the computational limitations of DiCE. Our method introduces dropout training of the discriminator to promote sparsity, a diversity term to maximize distances among generated counterfactuals, and a masking method to handle immutable features, thereby ensuring plausiblity. We evaluate our method on two benchmarks and apply it to a breast cancer dataset as the case study. We compare their performance with DiCE and CGAN. The results demonstrate that SCGAN generates sparse, diverse, and plausible counterfactual instances, making it a valuable tool for understanding the causal relationships between pathologic response to NST and ICM features. By performing counterfactual analysis, we identified cause-and-effect relationships between breast MR imaging phenotypes, molecular features, and pathologic response to NST.

Further research is required to improve the efficiency and scalability of the method without retraining the model and better methods for handling categorical features. Tuning the coefficients of the regularization terms is a potential direction for future research to balance prediction probability and sparsity in generating counterfactual instances.

## Data Availability

All data produced in the present study are available upon reasonable request to the authors.

## Acknowledgments

The authors are grateful for the kind support provided by NIBIB of the National Institutes of Health under award number R21EB033923-01, ASU-Mayo Clinic Seed Grant, the Gerstner Family and the Brandt Young Scholarship from the Centers of Individualized Medicine and ASU-Mayo Clinic Summer residency program.

